# Human Kidney is a Target for Novel Severe Acute Respiratory Syndrome Coronavirus 2 (SARS-CoV-2) Infection

**DOI:** 10.1101/2020.03.04.20031120

**Authors:** Bo Diao, Chenhui Wang, Rongshuai Wang, Zeqing Feng, Yingjun Tan, Huiming Wang, Changsong Wang, Liang Liu, Ying Liu, Yueping Liu, Gang Wang, Zilin Yuan, Liang Ren, Yuzhang Wu, Yongwen Chen

**Affiliations:** Department of Medical Laboratory Center, General Hospital of Central Theater Command, Wuhan, Hubei province, 430015, People’s Republic of China; Institute of Immunology, PLA, Third Military Medical University, Chongqing, 400038, People’s Republic of China; Department of Forensic Medicine, Tongji Medical College, Huazhong University of Science and Technology, No. 13 Hangkong Road, Wuhan, Hubei province, 430030, People’s Republic of China; Department of Nephrology, Renmin Hospital of Wuhan University, Wuhan, Hubei province, 430060, People’s Republic of China; Department of Pathology, 989^th^ Hospital of PLA, Luoyang, 471000, Henan Province, People’s Republic of China; Hubei Chongxin Judicial Expertise Center, Wuhan, Hubei province, 430415, People’s Republic of China

**Author notes:** Corresponding author: **Yongwen Chen** (Ph. D.), Institute of Immunology, PLA, Third Military Medical University, Chongqing, 400038, People’s Republic of China. or **Yuzhang Wu** (Prof. & Chair). Equally to this work (Bo Diao, Chenhui Wang, Rongshuai Wang, Liang Ren, Yuzhang Wu, Yongwen Chen).

## Abstract

**BACKGROUND:** The outbreak of a novel coronavirus (SARS-CoV-2, previously provisionally named 2019 novel coronavirus or 2019-nCoV) since December 2019 in Wuhan, China, has become an emergency of major international concern. Apart from the respiratory system, it is unclear whether SARS-CoV-2 can also directly infect other tissues such as the kidney or induce acute renal failure.

**METHODS:** We conducted a retrospective analysis of estimated glomerular filtration rate (eGFR) along with other clinical parameters from 85 patients with laboratory-confirmed COVID-19 admitted to a hospital in Wuhan from January 17, 2020 to March 3, 2020. Kidney tissues from six patients with postmortem examinations were analyzed by Hematoxylin and Eosin (H&E) and *in situ* expression of viral nucleocaspid protein (NP) antigen, immune cell markers (CD8, CD68 and CD56) and the complement C5b-9 was detected by immunohistochemistry. Moreover, the viral particles in kidneys were also investigated by transmission electronic microscope (EM).

**RESULTS:** 27.06% (23/85) patients exhibited acute renal failure (ARF). The eldery patients and cases with comorbidities such as hypertension and heart failure more easily developed ARF (65.22% *vs* 24.19%, *p*< 0.001; 69.57% vs 11.29%, *p*< 0.001, respectively). H&E staining demonstrated kidney tissues from postmortems have severe acute tubular necrosis and lymphocyte infiltration. Immunohistochemistry showed that SARS-CoV-2 NP antigen was accumulated in kidney tubules. EM observation also demonstrated that viruses-like particles are visible in the kidneys. Viral infection not only induces CD68^+^ macrophages infiltrated into tubulointerstitium, but also enhances complement C5b-9 deposition on tubules.

**CONCLUSIONS:** SARS-CoV-2 induces ARF in COVID-19 patients. Viruses directly infect human kidney tubules to induce acute tubular damage. The viruses not only have direct cytotoxicity, but also initiate CD68^+^ macrophage together with complement C5b-9 deposition to mediate tubular pathogenesis.

## Introduction

In December 2019, a cluster of pneumonia cases caused by a novel severe acute respiratory syndrome coronavirus 2 (SARS-CoV-2) were reported in Wuhan, Hubei Province, China.^1-3^ This disease, now designated as coronavirus disease 2019 (COVID-19) by the WHO, rapidly spread to other cities of China and around the world.^**4-6**^ By March 3, 2020, 80270 cases were laboratory confirmed with 2981 deaths in China based on CCDC (Chinese Center For Disease Control And Prevention) reports.^7^

Some studies have suggested that SARS-CoV-2, SARS-CoV (Severe acute respiratory syndrome-Coronavirus) and MERS-CoV (Middle East Respiratory Syndrome-Coronavirus) share a common ancestor resembling the bat coronavirus HKU9-1.^8,9^ Recently, Shi *et al*., reported that SARS-CoV-2 interacts with human ACE2 (angiotensin converting enzyme-II) molecules *via* its Spike protein.^10^ In addition to respiratory organs, the expression of ACE2 protein has also been observed in human kidneys,^11,12^ and Cheng et al., reported that some COVID-19 patients had an elevated level of proteinuria.^13^ However, no evidence directly demonstrates that SARS-CoV-2 can infect the kidney to cause acute renal failure.

We retrospectively analyzed the clinical data on kidney function from 85 cases of COVID-19 who were admitted into the General Hospital of Central Theatre Command in Wuhan, Hubei Province, from January 17, 2020 to March 3, 2020. To elucidate the mechanism of renal injury, we also used H&E staining and immunohistochemistry to visually assess tissue damage and viral presence in kidney tissues from six biopsies.

## Methods

### Patients

Medical records from 85 COVID-19 patients (aged from 21 years to 92 years) with dynamic observation of renal function in General Hospital of Central Theatre Command in Wuhan from January 17, 2020 to March 3, 2020 were collected and retrospectively analyzed. Postmortem autopsies were conducted on six patients who had been admitted in Hospital. Diagnosis of COVID-19 was based on the New Coronavirus Pneumonia Prevention and Control Program (5^th^ edition) published by the National Health Commission of China. All the patients were laboratory-confirmed positives for SARS-CoV-2 by use of quantitative RT-PCR (qRT-PCR) on throat swab samples. This study was approved by the National Health Commission of China and Ethics Commission of General Hospital of Central Theatre Command ([2020]017-1) and Jinyintan Hospital (KY-2020-15.01). Written informed consent was waived by the Ethics Commission of the designated hospital for emerging infectious diseases.

### Data collection and definitions

We reviewed clinical records and laboratory findings for all the patients. All information was obtained and curated with a customized data collection form. eGFR was calculated by the CKD-EPI equation based on serum creatinine level, sex, race and age. Acute renal impairment was defined as a decline of eGFR by at least 30% of the baseline value on admission or below 90mL/min on admission. Three investigators (C Wang, Z Fen and Y Chen) independently reviewed the data collection forms to verify data accuracy.

### Tissue morphology detection and immunohistochemistry

Due to the special infection-control precaution of handling deceased subjects with COVID-19, postmortem examination was performed in a designated pathology laboratory. Kidney specimens from autopsy were retrieved for standard examination *via* light microscopy. Briefly, paraffin-embedded tissue blocks were cut into 3 μm slices and mounted onto poly-lysine-coated glass slides, and tissue injury was stained for by H&E. Sections were also used for immunohistochemistry as following: Antigen retrieval was performed by microwaving these sections in citrate buffer (10 mM, pH□6.0). The sections were then incubated in 3% BSA plus 0.1% H2O2 for 1h at RT to block nonspecific binding. The sections were then incubated overnight at 4 °C with primary anti-SARS-CoV-2 nucleocaspid protein (NP) antibodies (clone ID: 019, 1:100, rabbit IgG; Sino Biological, Beijing), anti-CD8 (Clone ID:4B11, 1:100, mouse IgG2b; BIO-RAD), anti-CD68 (Clone ID:KP1, 1:100, mouse IgG1; BIO-RAD), anti-CD56 (Clone ID:123C3, 1:100, mouse IgG1; BIO-RAD), anti-C5b-9 (clone ID: aE11, 1:100, mouse IgG; Dakocytomation) or rabbit-isotype antibody controls (1:100; Dako). Sections were further incubated with the HRP-anti-Rabbit secondary antibodies for 1h at RT. Peroxidase activity was visualized with the DAB Elite kit (K3465, DAKO), and the brown coloration of tissues represented positive staining as viewed by a light microscope (Zeiss Axioplan 2, Germany).

### Transmission electron microscopy

Two kidneys from 2 cases of autopsy were collected and fixed with 2.5% glutaraldehyde in phosphoric buffer (pH 7.4), post-fixed with 1% osmate, dehydrated with gradient alcohol, embedded in Epon 812, double-stained with uranium acetate and lead citromalic acid. The virus-like particles were observed under a JEM1200 transmission electron microscope (Jeol, Tokyo, Japan).

### Statistical analysis

Statistical analyses were performed using GraphPad Prism version 8.0 (GraphPad Software, Inc., San Diego, CA, USA). Categorical variables were expressed as numbers (%). p values are from χ2.

## Results

### 1. SARS-CoV-2 induces ARF in COVID-19 patients

We retrospectively analyzed laboratory kidney functions from 85 cases of COVID-19 patients, and focusing on eGFR. Results showed that 27.06% (23/85) COVID-19 patients exhibited acute renal failure. Patients who are elderly (≥60 years) or carry comorbidities (65.22% *vs* 24.19%, *p*<0.001, 69.57% *vs* 11.29%, *p*<0.001, respectively), such as hypertension (39.13% *vs* 12.90%, *p*=0.0007) and coronary heart disease (21.74% *vs* 4.84%, *p*=0.018) more easily developed ARF (**Table1**), suggesting renal function impairment is relatively common in COVID-19 patients.

**Table 1.**
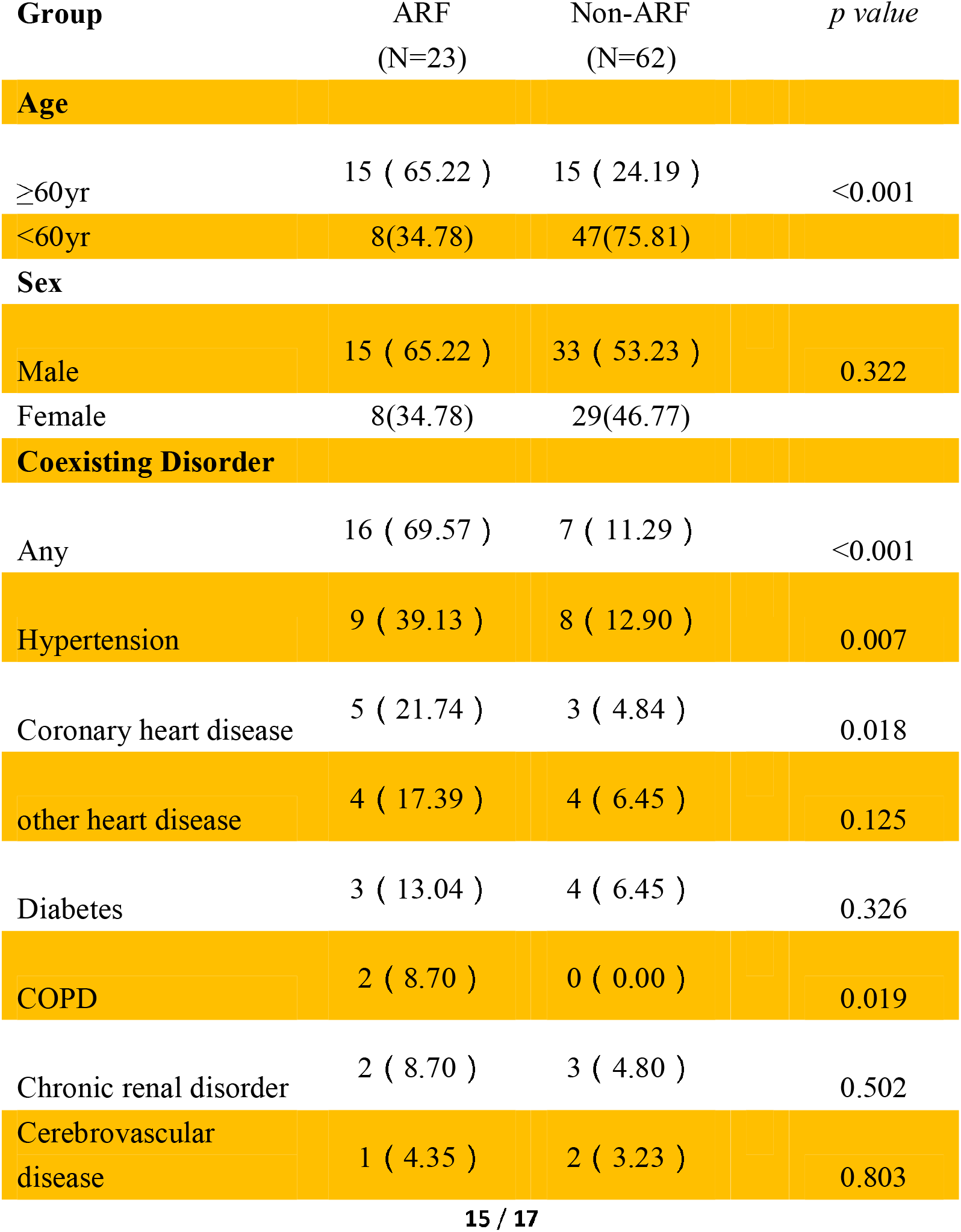
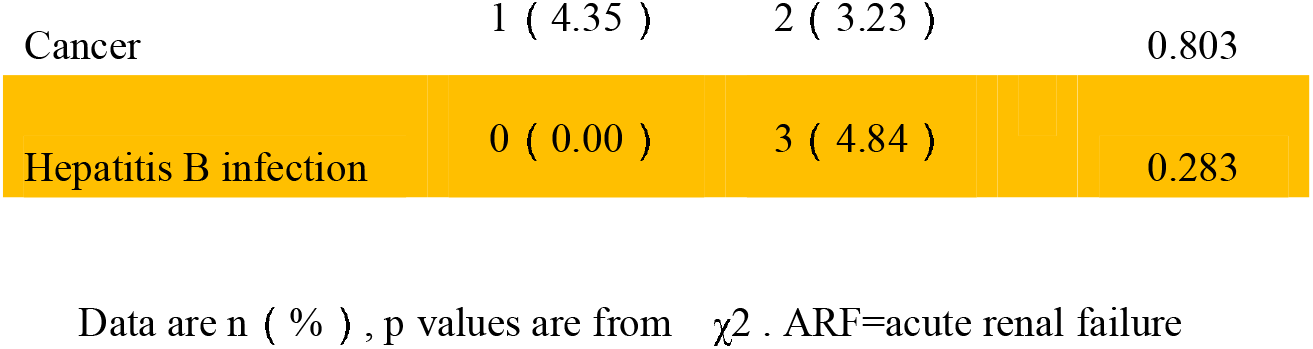
Clinical Characteristics of COVID-19 patients.

| Group | ARF<br>(N=23) | Non-ARF<br>(N=62) | *p value* |
| --- | --- | --- | --- |
| **Age** |  |  |  |
| ≥60yr | 15 ( 65.22 ) | 15 ( 24.19 ) | <0.001 |
| <60yr | 8(34.78) | 47(75.81) |  |
| **Sex** |  |  |  |
| Male | 15 ( 65.22 ) | 33 ( 53.23 ) | 0.322 |
| Female | 8(34.78) | 29(46.77) |  |
| **Coexisting Disorder** |  |  |  |
| Any | 16 ( 69.57 ) | 7 ( 11.29 ) | <0.001 |
| Hypertension | 9 ( 39.13 ) | 8 ( 12.90 ) | 0.007 |
| Coronary heart disease | 5 ( 21.74 ) | 3 ( 4.84 ) | 0.018 |
| other heart disease | 4 ( 17.39 ) | 4 ( 6.45 ) | 0.125 |
| Diabetes | 3 ( 13.04 ) | 4 ( 6.45 ) | 0.326 |
| COPD | 2 ( 8.70 ) | 0 ( 0.00 ) | 0.019 |
| Chronic renal disorder | 2 ( 8.70 ) | 3 ( 4.80 ) | 0.502 |
| Cerebrovascular disease | 1 ( 4.35 ) | 2 ( 3.23 ) | 0.803 |
| Cancer | 1 ( 4.35 ) | 2 ( 3.23 ) | 0.803 |
| Hepatitis B infection | 0 ( 0.00 ) | 3 ( 4.84 ) | 0.283 |

Data are n ( % ) , p values are from $\chi^2$ . ARF=acute renal failure

### 2. SARS-CoV-2 induces human kidney tubular injury

Next, the histopathological examination was performed by H&E-staining in kidney specimens from autopsy of six COVID-19 subjects with observed renal function impairment. Varying degrees of acute tubular necrosis, luminal brush border sloughing and vacuole degeneration, were found in different areas of all six renal specimens. Severe infiltration of lymphocytes in the tubulointerstitium was observed in two patients, and moderate infiltration was seen in three cases, and the remaining one case manifested absence of lymphocyte infiltration. Moreover, viral infection associated-syncytia were also observed in three cases. At the same time, dilated capillary vessels were observed in the glomeruli of these 6 cases. Nevertheless, severe glomerular injury was absent in all of these six cases, although benign hypertensive glomerulosclerosis and autolysis were noted in three hypertensive patients (**Figure 1, Table 2**). Collectively, these results demonstrated that SARS-CoV-2 infection mainly induces severe acute tubular necrosis and lymphocyte infiltration.

**Table 2.** The pathologic findings and protein expression of kidney tissues from COVID-19 patients undergoing postmortem examintion.

| Case number | Gender | Age | Coexisting disorder | H&E staining |  |  |  | Immunohistochemistry |  |  |  |  | SARS-CoV-2 confirmed by PCR |
| --- | --- | --- | --- | --- | --- | --- | --- | --- | --- | --- | --- | --- | --- |
|  |  |  |  | Benign glomerulosclerosis | Tubular necrosis | Intestinal infiltration | Autolysis | Viral NP antigen | C5b-9 on tubules | CD68 | CD8 | CD56 |  |
| 1 | male | 70 | Hypertension >10 years | + | +++ | +++ | ++ | +++ | ++ | ++ | - | - | Yes |
| 2 | female | 86 | Hypertension >10 years; coronary disease >10 year | + | +++ | +++ | + | ++++ | ++++ | ++++ | ++ | + | Yes |
| 3 | male | 62 | None | - | ++ | ++ | + | +++ | ++ | +++ | ++ | + | Yes |
| 4 | male | 51 | Hypertension >10 years | + | ++ | ++ | + | +++ | + | ++ | - | +/- | Yes |
| 5 | male | 78 | Hypertension >5 years; Gout | + | ++ | ++ | + | +++ | ++ | ++ | +- | - | Yes |
| 6 | female | 67 | None | - | ++++ | - | + | +++ | + | +- | - | - | Yes |

**Figure 1.**
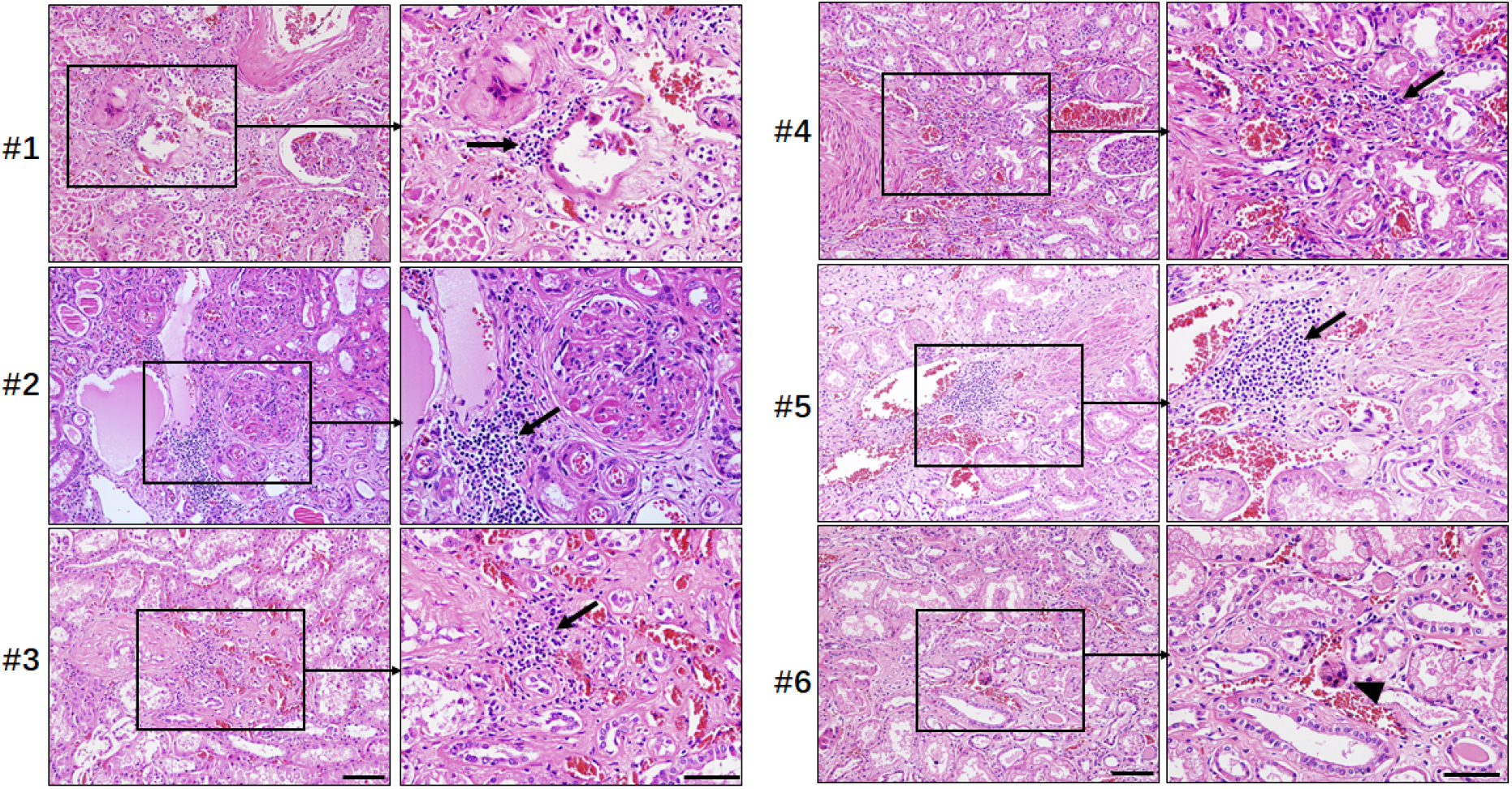
Representative H&E staining of kidney tissues from 6 cases of COVID-19 patients undergoing postmortem examination. Sections were stained by H&E, arrow indicated infiltrated lymphocytes; arrow head indicated viral infection associated-syncytia. Scale bar= 100 μM.

### 3. SARS-CoV-2 directly infects human kidney tubules

To confirm that SARS-CoV-2 might directly infect kidney tubules thus lead to tubular damage, the viral NP antigen was assessed by immunohistochemistry. Results showed that SARS-CoV-2 NP antigens could be seen in kidney tissues from all of these six samples, with NP expression restricted to kidney tubules, and NP-positive inclusion body was also observed. The NP-positive expression was observed in cytoplasm, but nuclear signals were not observed **(Figure 2A)**. Some NP-positive tubular epithelial cells were dropped from normal tubules (**Figure 2B**), while NP-antigen was also absent in the glomerulus (**Figure 2C**). The expression of NP protein in lung tissues from COVID-19 patients was used as positive controls (**Figure 2D**), whereas normal kidney tissues (**Figure 2E**) or renal sections from unrelated autopsies treated with rabbit isotype control antibodies (**Figure 2F**) were used as negative controls. Collectively, these data suggest that SARS-CoV-2 directly infects kidney tubules.

**Figure 2.**
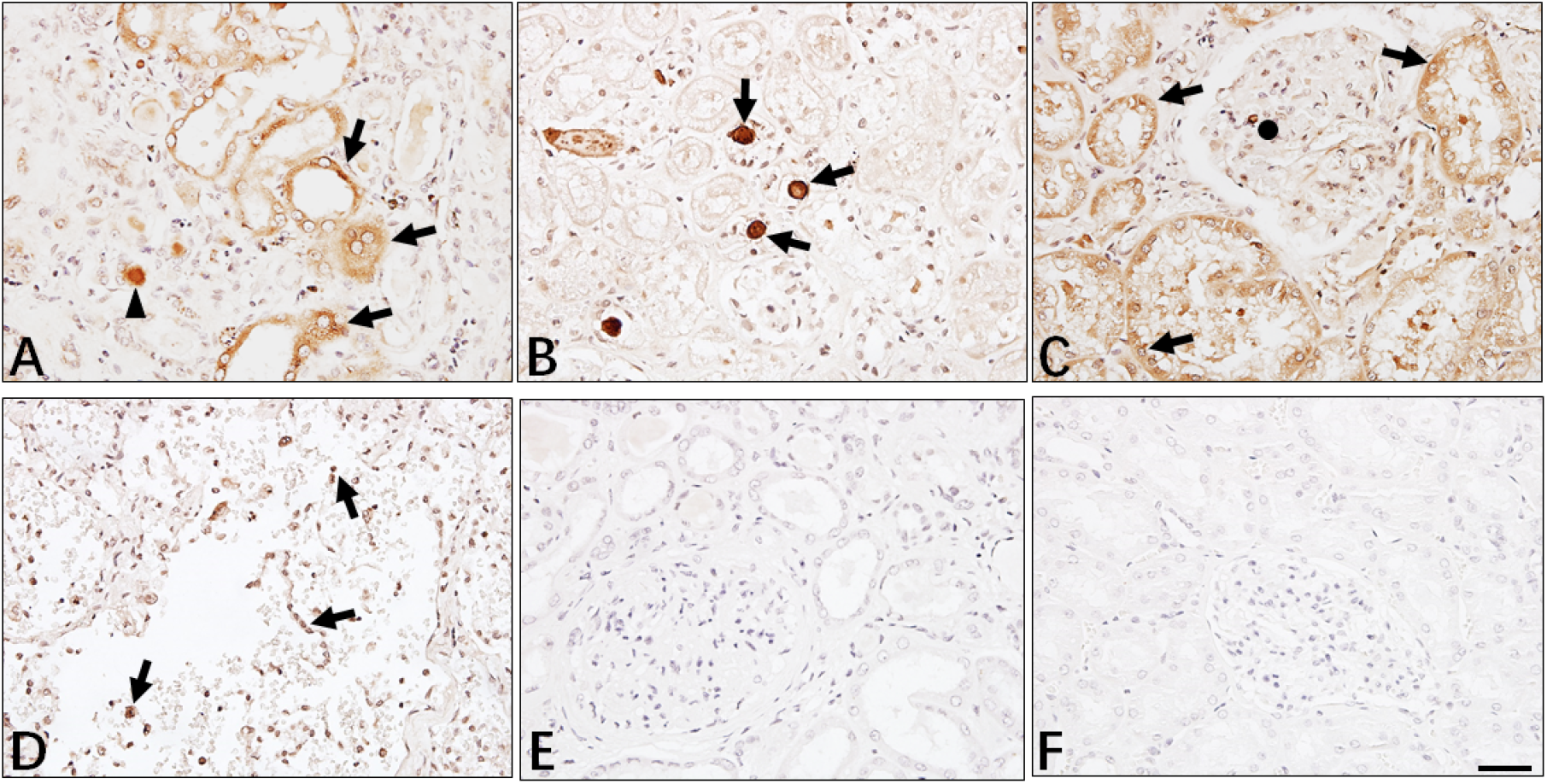
Immunohistochemistry analyzed SARS-CoV-2 NP antigen in kidney tissues. The expression of viral NP antigen was detected in kidney (**A**∼**C**) and lung (**D**) tissues from COVID-19 patients undergoing postmortem examination. **A**: arrow indicated NP positive tubules and arrow head indicated viral inclusion body. **B**: arrow indicated viral NP positive cells dropped from normal tubule; **C**: arrow indicated NP positive cells and circle indicated glomerulus; **E:** Absence expression of viral NP protein in normal kidney tissues; **F:** Sections incubated with rabbit IgG1 isotype control antibodies. Scale bar= 100 μM.

### 4. Direct observation of virus-like particles in kidneys by EM

Transmission electron microscopy (EM) was used to detect virions and virus-like particles in two kidneys from two autopsies. The results showed that cells in infected renal tissues were markedly swollen with expansion of mitochondria and lysosomes. The rough endoplasmic reticulum (RER) and smooth endoplasmic reticulum (SER) are also dilated greatly. Interestingly, viruses-like particles with 80∼160 nm diameters are observed in the broken lysosomes in cytoplasm, clearly identifiable due to high electron density and complete coating (**Figure 3**). Moreover, virus-like particles are all seen in both cases, demonstrating SARS-CoV-2 directly infects kidneys.

**Figure 3.**
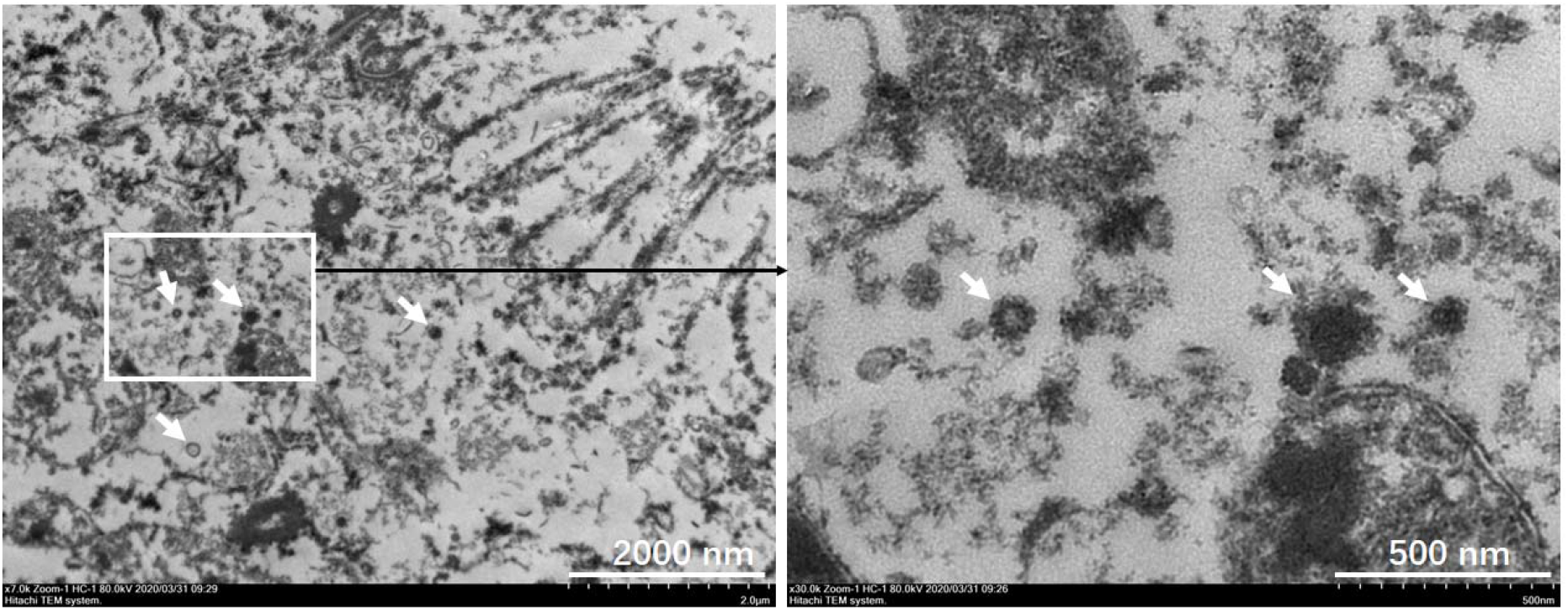
The virus-like particles are observed in kidney by EM. EM was used to detect virions in two kidney tissues from two autopsies, and viruses-like particles with 80∼160 nm diameters are observed in the broken lysosomes in cytoplasm, clearly identifiable due to high electron density and complete coating. Arrow indicated virus-like particles.

### 5. SARS-CoV-2 induces CD68^+^ macrophage infiltration and complement C5b-9 deposition

Since the infiltration of proinflammatory cells can significantly accelerate tubular damage, we next examined the identity of these infiltrated cells. Immunohistochemistry showed strong presence of CD68^+^ macrophages were in the tubulointerstitium of these six cases, while moderate numbers of CD8^+^ T cells were also observed in two cases, whereas, CD4^+^ T cells and CD56^+^ natural killer (NK) cells were seldom found in the examined tissues (**Figure 4A, Table 2**), suggesting that SARS-CoV-2 might cause further tubular damage through recruiting macrophages to infiltrate into the tubulointerstitium.

**Figure 4.**
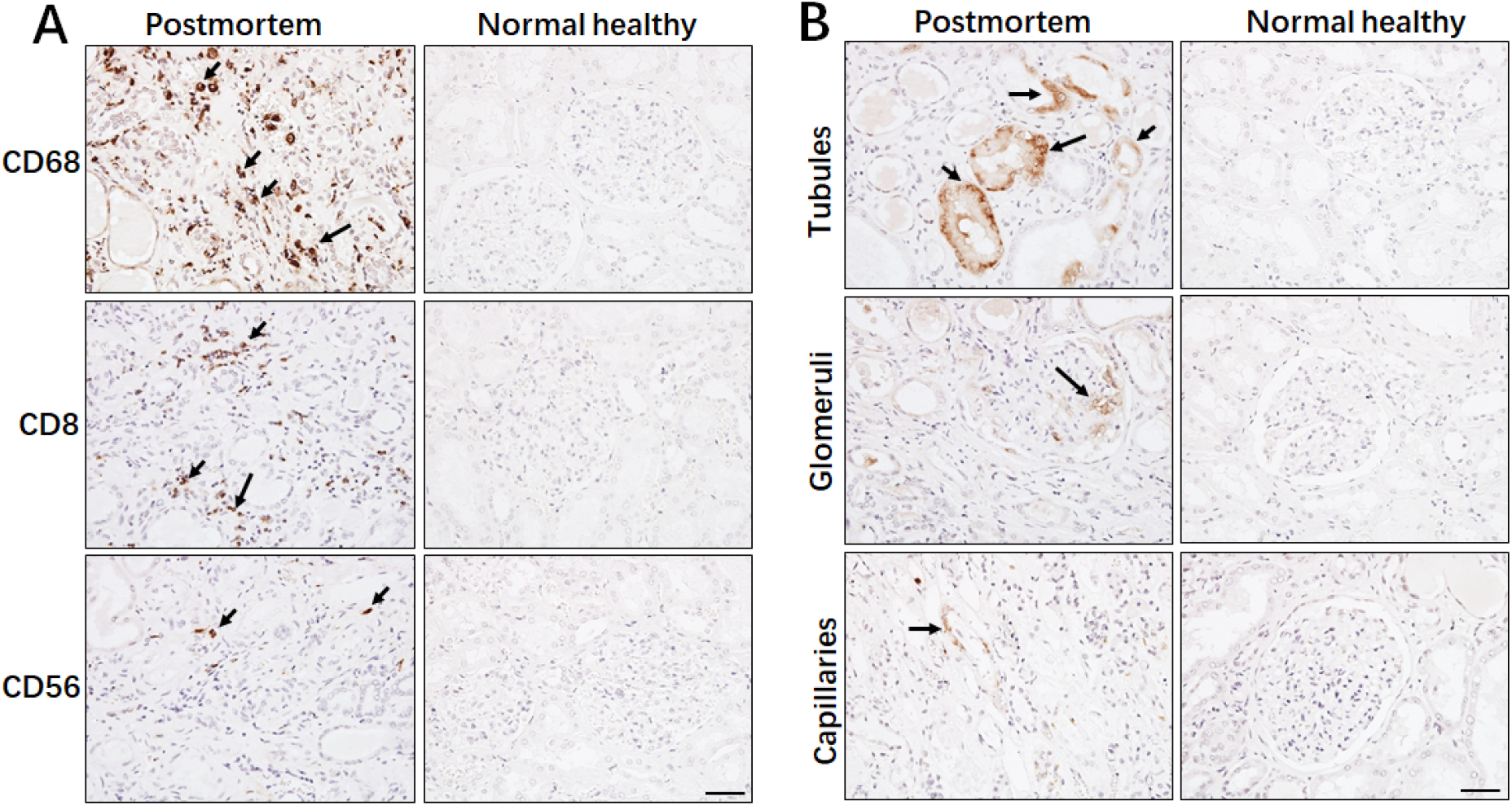
Immunohistochemistry analyzed lymphocytes and the complement C5b-9 in kidney tissues. The expression of **(A)** CD68, CD8 and CD56; **(B)** C5b-9 in kidney tissues from COVID-19 patients undergoing postmortem examination or normal healthy was detected by immunohistochemistry. Arrow indicated positive cells. Scale bar= 100 μM.

As the deposition of the complement membrane-attack complex (MAC, also named C5b-9) on tubules or glomeruli may also cause renal damage,^14^ we further analyzed C5b-9 status in kidney tissues from these postmortems. Interestingly, strong C5b-9 depositions on tubules were observed in all of these six cases. Very low levels of C5b-9 deposition on glomeruli and capillaries were also seen in two cases. However, C5b-9 expression is absent in normal kidney tissue (**Figure 4B, Table 2**), suggesting that SARS-CoV-2 infection induces acute tubular necrosis and ARF through triggering C5b-9 deposition.

## Discussion

In this study, we found that 27.06% of patients with COVID-9 had abnormal eGFR, and patients who are aged or have comorbidities more commonly developed ARF (**Table 1**), suggesting that ARF is relatively common following SARS-CoV-2 infection. This phenomenon is different from the 2003 outbreak of SARS, in which ARF was uncommon, despite being one of the top risk factors for mortality.^15^ Recently, Yan et al., reported that 63% (32/51) of COVID-19 patients had an elevated level of proteinuria.^13^ Collectively, these results illustrate that SARS-CoV-2 mediated ARF may be one of the major causes of multiorgan failure and eventual death in COVID-19 patients.

The extents of tubular atrophy and interstitial disease are the strongest histological parameters which correlate with renal function and predict progression. To further analyze how SARS-CoV-2 affects renal function, we focused on analyzing renal tissue morphology from autopsies. H&E staining showed that acute renal tubular damage and lymphocyte infiltration was observed in all six cases during postmortem examinations, while the glomeruli were intact, except for some cases with slight glomerulosclerosis, suggesting that other conditions such as hypertension and diabetic nephropathy may have been involved in the pathogenesis(**Figure 1)**. It has been shown that the ACE-2 receptor of SARS-CoV-2 is highly expressed in renal tubules.^12,16^ We used immunohistochemistry to analyze the expression of viral NP antigen *in situ* in kidney tissues, and found that viral NP antigen was restricted to the renal tubular cells of the infected tissues (**Figure 2**). Most important, EM observation demonstrated that viruses-like particles are also visible in the infected kidneys (**Figure 3**). This phenomenon is consistent with SARS-CoV and MERS-CoV, both of which can also infect human kidneys.^17,18^ Recently, some researchers have reported they have successfully isolated SARS-CoV-2 virus particles from the urine of COVID-19 patients, suggesting that kidney-originated viral particles may enter the urine through glomerular filtration. Taken together with our study, these results demonstrate that the kidney is also a site of viral infection and replication outside of the lungs.

While SARS-CoV-2 is a cytopathic virus which can also directly induce renal tubular injury during infection and replication, the occurrence of such injury may also initiate complex immune responses. After all, although host immune cells can infiltrate into infected tissue to counteract viral replication, hyperactivation of immune cells may instead promote fibrosis, induce epithelial cell apoptosis, and cause microvasculature changes.^19-21^ We report here that SARS-CoV-2 virus recruits high levels of CD68^+^ macrophages to infiltrate into the tubulointerstitium (**Figure 4A**), suggesting that proinflammatory cytokines derived from macrophages would induce tubular damage. Both clinical and experimental models suggest that the abnormal presence of serum-derived complement components in the tubular lumen leads to the assembly of the complement C5b-9 (*via* the alternative pathway) on the apical brush border of tubular epithelial cells (TECs), and that this is an important factor in the pathogenesis of tubulointerstitial damage.^22-24^ We here observed that SARS-CoV-2 initiates complement C5b-9 assembly and deposition on tubules (**Figure 4B**). We therefore demonstrate that SARS-CoV-2 causes tubular damage through direct cytotoxicity, but also initiates immune-mediates tubule pathogenesis.

In conclusion, we have demonstrated that the SARS-CoV-2 virus can directly infect human renal tubules and consequently lead to acute renal tubular injury. Moreover, improvement of eGFR would increase the survival of COVID-19 patients who have ARF. We strongly suggest that applying potential interventions including continuous renal replacement therapies (CRRT) for protecting kidney function in COVID-19 patients, particularly for ARF cases, maybe a key method to preventing fatality.

## Data Availability

For protection of patients' privacy, all data used during the study are only be provided with anonymized version.

## Authors’ contributions

Yuzhang Wu, and Yongwen Chen were involved in the final development of the project and manuscript preparation; Zilin Yuan, Chenghui Wang and Zeqing Feng analyzed the data; Chenghui Wang did EM experiments; Bo Diao and Huiming Wang performed most of experiments; Yin Liu, Gang Wang, Yinjun Tan and Yueping Liu did H&E staining and immunohistochemistry; Changsong Wang evaluated H&E and immunohistochemistry results; Liang Liu, Rongshuai Wang and Liang Ren provided autopsies and analyzed H&E staining.

## Ethics committee approval

This study was approved by the National Health Commission of China and Ethics Commission of General Hospital of Central Theatre Command and Jinyintan Hospital.

